# Social interventions can lower COVID-19 deaths in middle-income countries

**DOI:** 10.1101/2020.04.16.20063727

**Authors:** Angel Paternina-Caicedo, Marc Choisy, Christian Garcia-Calavaro, Guido España, José Rojas-Suarez, Carmelo Dueñas, Adrian Smith, Fernando De la Hoz-Restrepo

## Abstract

A novel pandemic coronavirus disease (COVID-19) was first detected in late 2019 in Wuhan (China)^1,2^. COVID-19 has caused 77 national governments worldwide to impose a lockdown in part or all their countries, as of April 4, 2020^3^. The United States and the United Kingdom estimated the effectiveness of non-pharmaceutical interventions to reduce COVID-19 deaths, but there is less evidence to support choice of control measures in middle-income countries^4^. We used Colombia, an upper-middle income country, as a case-study to assess the effect of social interventions to suppress or mitigate the COVID-19 pandemic. Here we show that a combination of social distancing interventions, triggered by critical care admissions, can suppress and mitigate the peak of COVID-19, resulting in less critical care use, hospitalizations, and deaths. We found, through a mathematical simulation model, that a one-time social intervention may delay the number of critical care admissions and deaths related to the COVID-19 pandemic. However, a series of social interventions (social and work distance and school closures) over a period of a year can reduce the expected burden of COVID-19, however, these interventions imply long periods of lockdown. Colombia would prevent up to 97% of COVID-19 deaths using these triggered series of interventions during the first year. Our analyses could be used by decision-makers from other middle-income countries with similar demographics and contact patterns to Colombia to reduce COVID-19 critical care admissions and deaths in their jurisdictions.

---

The novel pandemic coronavirus disease (COVID-19), caused by SARS-CoV-2, first appeared in Wuhan, China, in late 2019^1,2^. As of April 4, 2020, this pandemic has resulted in 1,192,028 confirmed cases and 64,316 deaths worldwide^5^. COVID-19 has caused 77 national governments around the world to impose a lockdown in part or all their countries, as of April 4, 2020^3^. However, most world jurisdictions have not achieved the containment of COVID-19. Only China Hong Kong, Singapore, and South Korea had appeared to be successful so far in the suppression of transmission of COVID-19. China contained the virus after social isolation measures. Despite this, a report^6^ suggested that if these social interventions (non-pharmacological interventions) had been conducted one week, two weeks, or three weeks earlier in mainland China, cases in this region could had been reduced by 66%, 86%, and 95%, respectively. However, if these social interventions were instead conducted one week, two weeks, or three weeks later, the number of cases could have shown a 3-fold, 7-fold, and 18-fold increase, respectively. South Korea, instead, began systematically testing for the virus and isolating individuals reported to have COVID-19. Despite the success of South Korea in containing the virus, this approach may be difficult to implement in countries who lack comparable human, technological, and economical resources to support widespread case detection, contact tracing and testing.

Several social interventions have been proposed for those countries who fail to act early in the pandemic. These interventions may suppress (lower the effective reproductive value to less than one) or mitigate (reduce the epidemic peak) the number of cases, deaths, and other adverse outcomes of COVID-19. Some of these social non-pharmacological interventions include social distance, work distance, or closure of schools and universities. Imperial College London suggested the United Kingdom could expect 510 thousand deaths without suppression or mitigation, and a reduction between 92-98% of deaths with several social interventions^4^. The report by Ferguson et al. (2020)^4^ simulated an intermittent intervention that would be triggered in the United Kingdom by reaching between 60 and 400 critical care admissions in a day, under several transmission scenarios.

The Ferguson et al.^4^ report and derivate analysis in Europe^7^ represent a roadmap of potential interventions worldwide. These analyses were only applied to high-income countries and did not consider other potential interventions that could be implemented to suppress or mitigate the pandemic for the next two years. Such modelling approaches may be suitable for application to middle- and low-income countries, however the demographic distribution of younger, and therefore healthier overall population, may influence their results and interpretation.

Here we attempt to inform decision-making in developing jurisdictions, by showing some interventions are not predicted to reduce the attack rate or adverse outcomes and only delay the epidemic peak, while a more comprehensive set of interventions triggered throughout a year may reduce resource use and deaths in a middle-income country. We took Colombia, a South American upper-middle income country, as a case-study to measure the expected number of deaths without and with social interventions. We therefore use an age-structured transmission dynamics model, in which the infected population goes through compartments of disease and can end either recover or die (**Extended Data Fig. 1**). This model was fitted and calibrated with empirical data (i.e. calibration targets) from Italy, Wuhan (China), the United States, and Colombia, using the following parameters:

1. The age-specific adjusted symptomatic case-fatality rate in China^8^.
2. The age-specific crude symptomatic case-fatality rate in Italy^9^.
3. The age-specific hospitalization rate given symptomatic in the United States^10^
4. The cumulative deaths in Italy^11^.
5. The cumulative number of critical care admissions in Italy^11^.
6. The cumulative number of hospital admissions in Italy^11^.
7. The cumulative cases in Wuhan (China)^8^.

We chose very specific data-points to analyze trends to include unsuppressed or unmitigated transmission dynamics (see Methods). These calibration targets were optimized using a bounded optimization algorithm for 200 starting values to obtain the 33 free parameters calibrated in the model: the age-specific probabilities of symptoms, hospitalization, critical care admission, and death while in critical care, along with the per capita transmission rate. We further used COVID-19 case data from Colombia to select the best set of parameters to fit this epidemiological model.

The resulting cumulative attack rate in the first 12 months in the calibrated model we formulated was 83%, for an R_0_ of 2.12, in line with previous empirical studies estimating the R_0_ of COVID-19 between 2 and 3^12,13^.

Colombia, a country of approximately 50 million people, had 1,406 cases and 32 deaths by April 4, 2020^14^. The best fitting parameters in calibration suggest the percentage of asymptomatic people in the entire age-distribution of Colombia is 73%. This percentage of asymptomatic is similar to previous studies using both mathematical models and Chinese empirical data on the asymptomatic percentage of COVID-19 in close contacts^15,16^. A total of 60% of the population older than 60 years in the present model were symptomatic (**Extended data Fig. 2**).

Under the fitted transmission parameters, Colombia can expect 288.8 deaths per 100,000 pop. without any social intervention, with 83% of all deaths occurring among people >60 years old (**Fig. 1**). This estimate does not include the likely additional mortality effect of overwhelmed healthcare facilities during the peak of the epidemic.

**Fig. 1.**
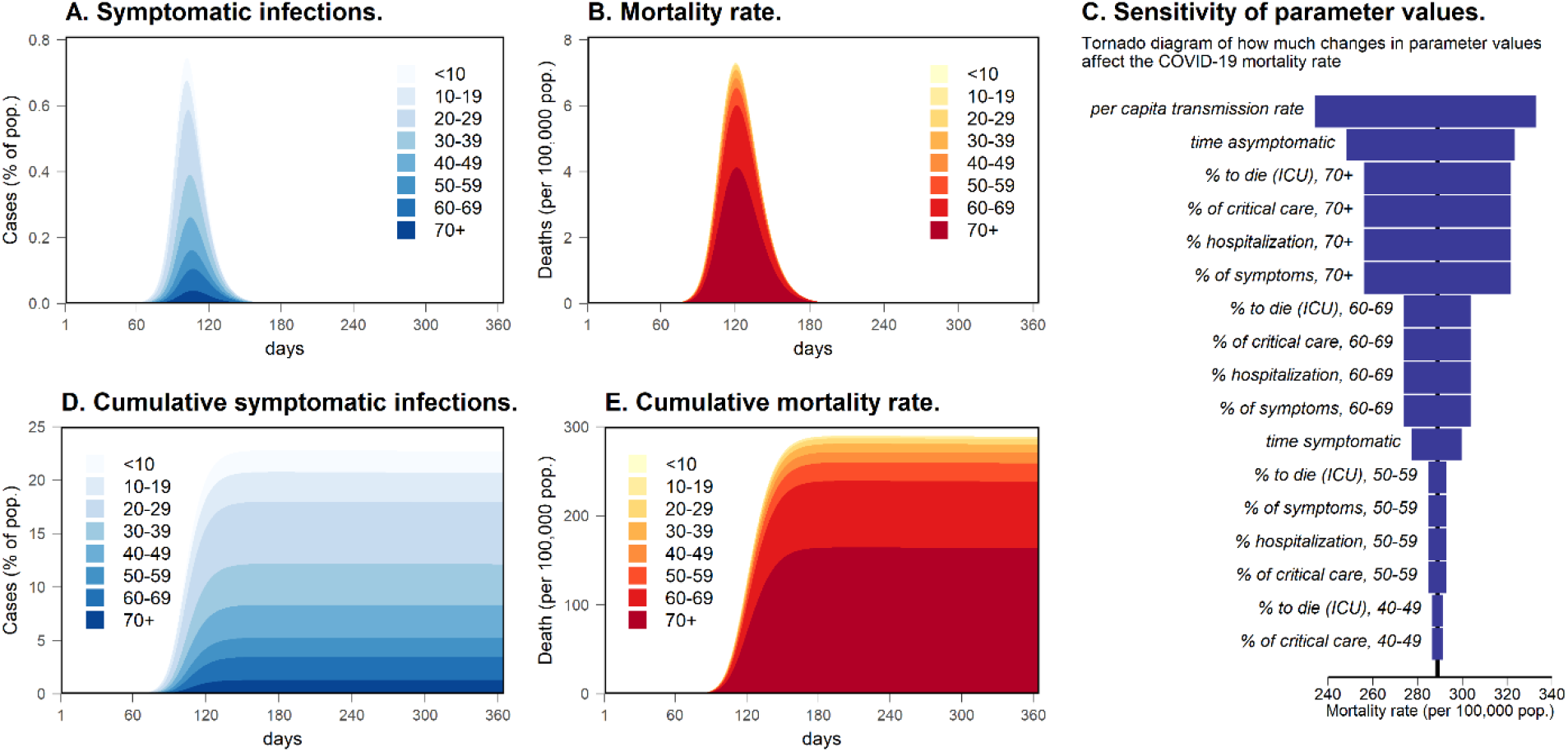
Scenario without social interventions to suppress or mitigate COVID-19 in Colombia. **Note**: Panel D and E show the cumulative rate for the population of Colombia. Colors in Panels A, B, D, and E show the symptomatic infections (A and D) and mortality (B and E) of each age-group over the population of the country.

## One-time social interventions will not permanently reduce transmission

The model predicts that a one-time social intervention lasting three or four months will not suppress or mitigate COVID-19 during a year, but will delay the epidemic peak for the time this one-time intervention is implemented. Our results suggest that applying social distance, work distance, and schools/universities closure for three or four months can suppress or mitigate transmission while implemented, but will not lead to elimination of COVID-19 transmission (**Table** and **Fig.1**). Whilst social interventions can better prepare the healthcare system for the pandemic, after lifting the intervention, we predict that COVID-19 will continue its spread in the population, producing a similar attack rate and deaths reduced between 0.3% and 0.7% (**Table** and **Fig. 2**).

**Table.** Dynamics of COVID-19 in Colombia after 365 days, with and without social interventions.

| Intervention | Attack rate | Symptomatic people reported to healthcare | Critical care admissions | Mortality |
| --- | --- | --- | --- | --- |
|  | (%) | (%) | *n (per 100,000 pop.)* | *per 100,000 pop. (% reduction)* |
| **No intervention** | 64.7 - 90.7 | 14.4 - 29.6 | 936,549 (1,824.1) | 288.8 |
| **Social interventions for 3 months** |  |  |  |  |
| School closures | 64.8 - 88.3 | 14.4 - 28.9 | 934,765 (1,820.6) | 288.0 (0.3) |
| Social distancing | 64.8 - 84.7 | 14.4 - 27.4 | 934,750 (1,820.6) | 287.9 (0.3) |
| Work distance | 64.7 - 89.3 | 14.4 - 29.1 | 926,975 (1,805.4) | 286.7 (0.7) |
| Social distancing and school closures | 64.8 - 77.0 | 14.4 - 23.7 | 934,191 (1,819.5) | 287.6 (0.4) |
| All interventions | 46.1 - 91.0 | 11.1 - 29.6 | 933,187 (1,817.6) | 286.9 (0.7) |
| **Intermittent interventions (triggered at 50 critical care admissions)** |  |  |  |  |
| School closures | 52.5 - 88.3 | 11.8 - 28.9 | 874,336 (1,702.9) | 269.7 (6.6) |
| Social distancing | 35.6 - 83.7 | 7.8 - 27.1 | 736,690 (1,434.9) | 223.4 (22.7) |
| Work distance | 60.8 - 89.3 | 13.4 - 29.1 | 900,821 (1,754.5) | 281.1 (2.7) |
| Social distancing and school closures | 22.3 - 69.2 | 4.6 - 21.0 | 300,254 (584.8) | 58.0 (79.9) |
| All interventions | 6.2 - 35.8 | 1.3 - 11.3 | 43,915 (85.5) | 8.4 (97.1) |
| **Intermittent interventions (triggered at 100 critical care admissions)** |  |  |  |  |
| School closures | 52.7 - 88.3 | 11.9 - 28.9 | 875,188 (1,704.6) | 270.0 (6.5) |
| Social distancing | 36.4 - 83.7 | 8.0 - 27.1 | 739,422 (1,440.2) | 224.3 (22.4) |
| Work distance | 60.9 - 89.3 | 13.4 - 29.1 | 901,075 (1,755.0) | 281.1 (2.7) |
| Social distancing and school closures | 24.3 - 69.3 | 5.0 - 21.0 | 313,082 (609.8) | 62.1 (78.5) |
| All interventions | 9.9 - 36.7 | 2.1 - 11.6 | 101,303 (197.3) | 21.2 (92.7) |

**Fig. 2.**
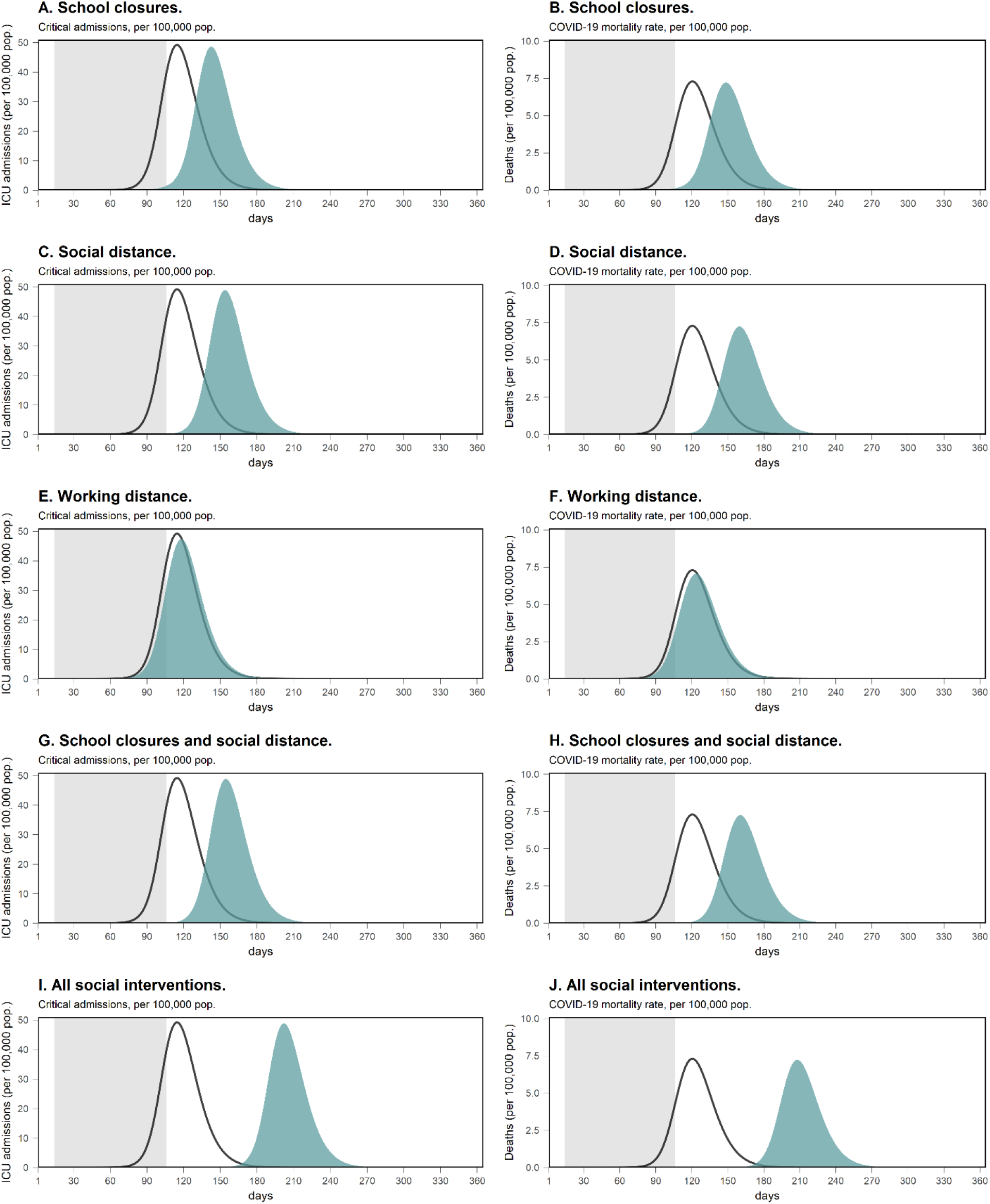
Critical care admissions and deaths related to COVID-19 dynamics with a three-month social intervention in Colombia, according to the type of intervention (gray area) compared to no intervention (black line).

## Intermittent suppression and mitigation will reduce up to 97% of deaths

We simulated a similar scenario to the report of Ferguson et al.^4^ We used triggers of action, in which achieving 50, 100, or 150 daily admissions to critical care in Colombia will trigger a social intervention for a week, after which the intervention is evaluated every week until the number of critical care admissions is above the trigger.

We estimate that a combination of school closures, social distancing, and work distancing may reduce deaths by 97% with a trigger of action of 50 critical care admission in a day. This combination of interventions would have to be active for at least 81% of the year (around 10 months) (**Extended Data Fig. 3**). A trigger of 100 critical admissions with all these interventions would reduce deaths by 93%, while this reduction is 92% for 150 admissions, and 200 admissions the death reduction was 90%.

A combination of social interventions after a trigger of 50 critical admissions will decrease the cumulative infection attack rate in a year from 83% to a range between 6-36%. (**Table, Fig. 3**, and **Extended Data Fig. 3**). We predict that social distancing alone, will result in one peak, and will reduce the COVID-19 mortality rate by 23% (**Table, Fig. 3**, and **Extended Data Fig. 4**).

**Fig. 3.**
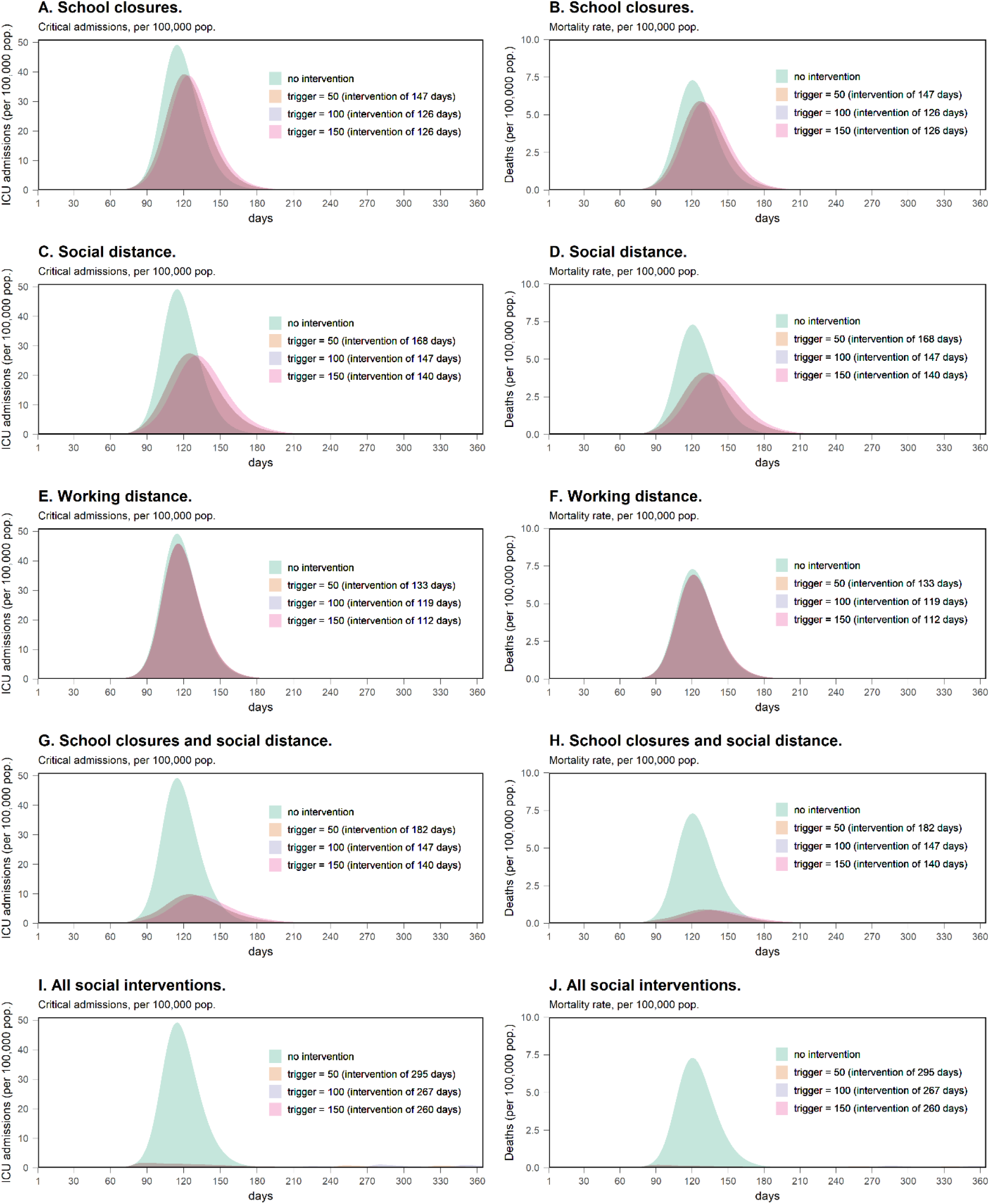
Critical care admissions and deaths related to COVID-19 dynamics with intermittent social interventions for a year in Colombia, triggered by 50, 100, or 150 number of daily admissions to critical care.

We also estimate that the effective reproductive number will be reduced to less than 1 after 70 days since the start of the COVID-19 epidemic, or within a week since starting all the social non-pharmacological interventions, and with a trigger of 50 critical care admissions in a day (**Extended Data Fig. 5**). Using a trigger of 100 critical care admissions in a day would result in a lower effective reproductive number to less than 1 after 77 days, and within a week of starting the home, work, school, and social distancing.

## Limitations

Our analysis is a mathematical simulation containing several assumptions (see Methods). In addition, our fitting is as good as the quality of the target data from China, the United States, and Italy we used to calibrate the simulation. Our model assumptions reflect key parameters for which there is not enough good quality data. One of the most important parameters changing the overall results in sensitivity analysis was the number of days an asymptomatic person is infectious. We assumed five days as the length of this period. In **Panel C** of **Fig. 1** the asymptomatic infectious time is the second parameter that would change results the most, and for which there is not much empirical data. Other parameters calibrated to data result in an epidemiological profile for COVID-19 in the model compared to empirical data of China, Italy, and the United States (**Extended data Fig. 2**). We also assume immunity after COVID-19 infection for the length of the simulation (18 months in **Extended data Fig. 3** and **Extended data Fig. 4**).

Our calibration targets come from countries with a different age-structure compared to Colombia, an upper-middle income country. While some of our calibration targets are age-stratified, the cumulative trend of hospitalizations, critical care admissions, and deaths in Italy is an aggregate of the population, potentially increasing the curve of severe cases in the base model scenario. This also means that parameters from China, Italy, and the United States are less robust in middle-aged population, producing more uncertainty in our estimations.

For low- and middle-income countries, the need of good quality empirical data is important to increase the level of evidence in the decision-making process, good quality data is needed as the pandemic spreads worldwide.

## Conclusions

Although our analyses estimate the expected burden of COVID-19 in a middle-income country with and without non-pharmacological social interventions, COVID-19 is still an evolving situation worldwide, and many uncertainties remain for this disease. Our analyses take the experience of China, Italy, and the United States, and attempt to provide a rough guide of interventions to prevent these adverse outcomes in an upper-middle income country like Colombia. The governments worldwide should consider economic and other societal externalities resulting from the implementation of these trigger thresholds and series of interventions.

Our analyses provide decision-makers from middle-income, and potentially in poor economies, with evidence and data to base their decisions. While the trigger of 50 critical care admissions was effective in Colombia to prevent deaths, the trigger value would depend on the available critical care units of each country.

## Data Availability

All data used for the present study is open data. Readers wishing to replicate the study can access the data under a reasonable request to the corresponding author.

## Disclaimer

The COVID-19 pandemic is an ongoing situation worldwide, and this is a preprint paper not yet peer-reviewed. Therefore, the results of this research may change, according to recommendations of other experts. Despite this, the authors in this research present this analysis to aid decision-making worldwide, confident in the interpretation of the main aim, showing that social non-pharmacological interventions can reduce COVID-19 burden in upper-middle income countries with younger and overall healthier populations.

## Methods

We developed a system of ordinary differential equations (ODE) to simulate the expected number of infections, cases, hospitalizations, critical care admissions, and deaths resulting from the COVID-19 pandemic.

### The ODE model

The ODE model follows the Colombian population in 16 age-groups (0-4, 5-9, 10-14, 15-19, 20-24, 25-29, 30-34, 35-39, 40-44, 45-49, 50-54, 55-59, 60-64, 65-69, 70-74, and 75+), through nine compartments: 1) susceptible; 2) exposed; 3) symptomatic infectious; 4) asymptomatic infectious; 5) complicated symptomatic (at homecare) infectious; 6) complicated hospitalized (isolated) not infectious; 7) complicated needing critical care (isolated) not infectious; 8) recovered from symptomatic infection; and 9) recovered from complicated infection.

The model follows a hypothetical cohort of an age-stratified population in Colombia every day for a year, assuming the population of 2020. When the susceptible population is exposed, persons can go to the symptomatic or asymptomatic compartment. If asymptomatic, the person is assumed to recover in the base scenario in five days; while if symptomatic, the population can go to home isolation and then recover, or go into hospitalization. A person hospitalized can either recover or go to the critical care unit. Finally, people in critical care can either die of COVID-19 or go to the recover compartment (**Extended Data Fig. 1**). The ODE mode includes births in the susceptible compartment and all-cause mortality in all compartments, except in critical care and the death compartments.

The ODE model uses Colombian age-specific contact rates from a previous study^17^ to fit the force of infection. All models and analyses were made in R (v3.6.3).

The ODE system is represented by the following equations:

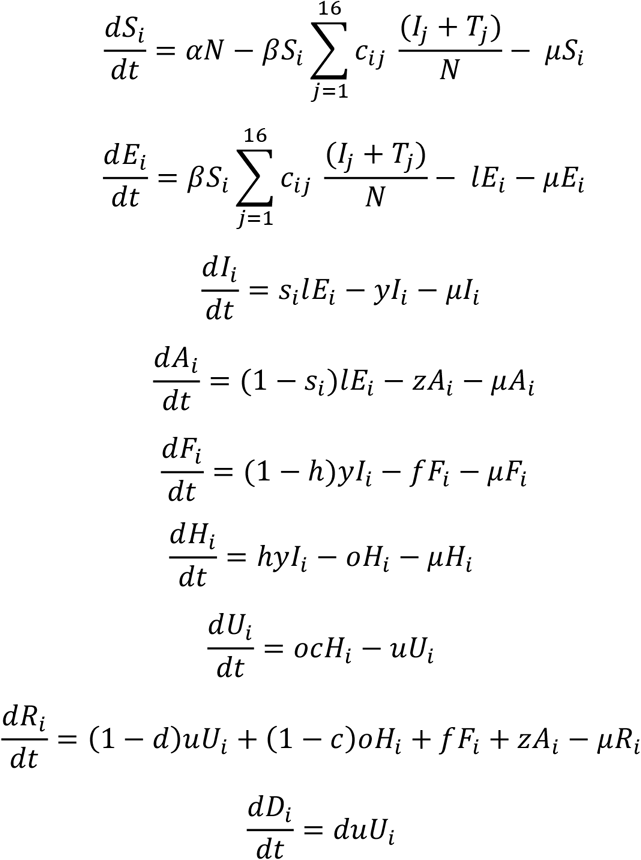

1. Susceptible (*S*).
2. Exposed (*E*).
3. Symptomatic infectious needing health care attention (*I*).
4. Asymptomatic infectious (*A*).
5. Complicated symptomatic (at homecare) not infectious (*F*).
6. Complicated hospitalized (isolated) not infectious (*H*).
7. Complicated needing critical care (isolated) not infectious (*U*).
8. Individuals recovered from infection (*R*).
9. Death (*D*).

Where the fixed parameters are: 1/*l* is the incubation time (5 days) ^13,18^, time in complicated symptomatic infectious (*f*) (assumed as 4 days), 1/*z* is the time while asymptomatic infectious (assumed as 5 days), 1/*y* is the time while symptomatic and infectious (assumed as 5 days), 1/*u* time in critical care (5 days)^13^, 1/*o* is time hospitalized (5 days)^13^, *µ* is the all-cause mortality rate in Colombia^19^, and *α* is the birth rate (13,2 per 1,000 pop.)^19^ All these values were parametrized per day. The free calibrated parameters were the per-capita transmission rate (β) (for all age-groups); and the age-specific probability of symptoms given infected (*s*), hospitalization probability (*h*), critical care probability (*c*), and death rate while in critical care (*d*) (in eight age-groups, 0-9, 10-19, 20-29, 30-39, 40-49, 50-59, 60-69, 70+). In the model, *c* is the contact rate per day, at age *i*, with a contact of age *j*.

The ODE model makes the following assumptions:

1. The infection with SARS-CoV-2 follows the pattern shown by the classical ODE susceptible-exposed-infected-recovered model.
2. The population follows the age-distribution of Colombia, and the contact rate we used to simulate contacts between age-groups is an accurate report of the contacts in this country.
3. The immunity of COVID-19 infectious lasts for 18 months at least (the length of the simulation in this report).
4. The incubation period lasts an average of five days, according to previous studies in China^13,18^.
5. The model assumes one index case starts the epidemic, and no other importation occurs for the length of the modeled time horizon.
6. Asymptomatic transmission is also assumed to occur in this model, as previously reported^20^.

### Calibration of the ODE model

The model was calibrated using the Hooke-Jeeves algorithm for derivative-free optimization. We used seven calibration targets plus a comparison with Colombian data, to match the dynamics of the ODE model. We used 200 starting values, randomly assigned using latin hypercube sampling. We compared observed versus predicted values with log-likelihoods using the binomial distribution, finding the best fitting set of targets for the country

The calibration of the ODE model used the following targets:

1. Age-specific adjusted symptomatic case-fatality rate by age in China^8^.
2. Age-specific crude symptomatic case-fatality rate by age in Italy^9^.
3. Age-specific hospitalization probability in the United States^10^. The data was in different age-groups for this calibration target, we therefore modeled the gaps of the percentage hospitalized between age-groups with a four-degree polynomial linear regression.
4. Cumulative deaths in Italy^11^, starting from the date of ten deaths until either 15 days have passed after first case detected or the lockdown in March 9, 2020
5. The cumulative number of critical care admissions in Italy^11^, starting from the date of 50 critical care admissions until either 15 days have passed after the first case detected or the start of the lockdown.
6. The cumulative number of hospital admissions in Italy^11^. Starting from the date of 150 hospitalizations until either 15 days have passed after the first case detected or the start of the lockdown.
7. Cumulative cases in Wuhan (China)^8^, From the date ten cumulative cases are detected, until either 45 days have passed or the start of the lockdown in Wuhan in January 23, 2020.

Each of these 200 set of parameters, from the same number of starting values, was fitted again to calculate the difference between simulated and observed case data in Colombia before the lockdown on March 20 of 2020^14^ (**Extended data Fig. 6**).

### Calculation of R_0_

We approximated the R_0_ by using the formula by Dietz (1993)^21^:

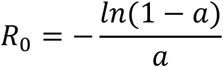

Where *a* is the attack rate. This formula assumes homogeneous mixing and a closed population. The R_0_ for our calibrated model was 2.12, according to the approximation using the attack rate method.

We also included an estimation of the effective reproductive value (R_t_), according to formulas by Cori et al. (2013)^22^, used in the R package EpiEstim (**Extended Data Fig. 5**).

### Contact matrices and simulated interventions

We used the Prem et al. (2017) estimation of contact matrices for Colombia^17^. These contact matrices are composed of four matrices (home, work, school, and other contacts).

We simulated five interventions with these matrices:

– School closures: We changed all school contacts to zero.
– Social distance: We assumed the matrix of other contacts are reduced by 80% (contacts other than home, work or work).
– School closures and social distance: We simulated the outcomes of reducing work contact by 70% and increase contacts of individuals of the same age (50% for persons <20 years of age and 10% for the other age-groups), as reported by Prem et al.^17^
– Work distancing: We assumed 80% of the working population would reduce a 50% their contact during work.

For the triggered interventions, we assumed the data on critical care admissions was evaluated on the fifth day of each week. Each of these social intervention lasts one week, where it was evaluated again every week until the critical care admissions per day were lower than the selected trigger.

## Code availability

The models were made with R (v3.6.2), using the deSolve package (v1.28) with compiled code in C. The C code for the function to run the ODE model is shown in the Supplementary Appendix. We ran the analysis in a Windows 10 computer, Core i9 of 8^th^ generation of 6C/12T. Each of the ODE model simulations ran in ∼0.5 seconds. We additionally used the packages EPiEstim (v2.2-1), dfoptim (v2018.2-1), matrixStats (v0.5.5.0), tidyverse (v1.3.0) scales (1.0.0), gridExtra (v2.3), parallel (v3.6.2), foreach (v1.4.7), doParallel (v1.0.15), and foreign (v.0.8-72). The entire code for replicability can be requested to the corresponding author and will be published in www.covid19app.care.

## Author Contributions

A.P.-C. helped design, analyze, and write the first draft of the study. M.C. helped analyze and interpret the study, and provided critical intellectual content. G.E. helped design, analyze, and interpret the study, and provided critical intellectual content. C.G.-C. helped design, analyze, and write the first draft. J.R.-S., C.D., and A.S. helped analyze and interpret the study, and provided critical intellectual content. F.D.-R. helped design, interpret, and analyze the study. All authors approved the final version of the manuscript.

---

**Extended data Fig. 1.**
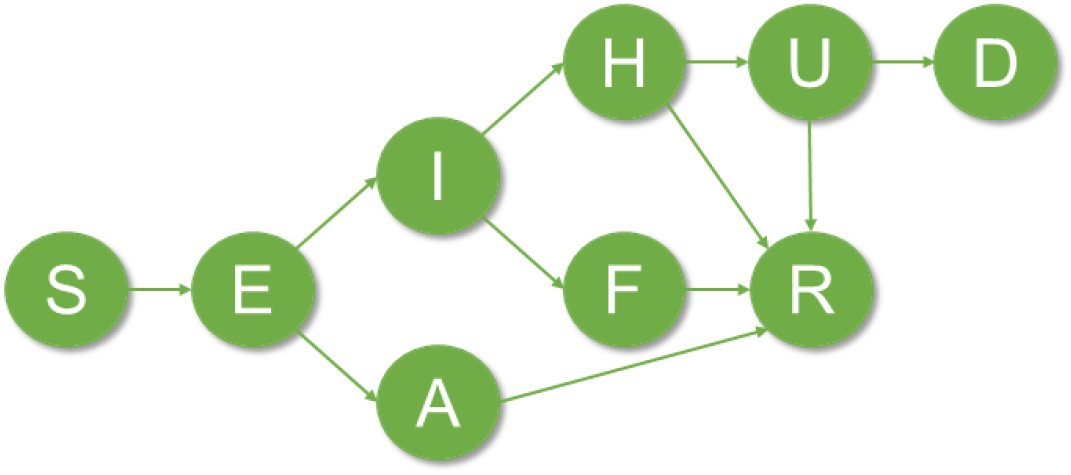
Transmission model of COVID-19, using compartments in a system of ordinary differential equations. **Note**: The model follows an age-stratified hypothetical population from Colombia infected with COVID-19, through nine compartments: susceptible (S), exposed (E), infectious symptomatic (I), infectious asymptomatic not needing medical care (A), hospitalized (isolated) (H), symptomatic at home (isolated) (F), severe in critical care (isolated) (U), recovered (R), and deceased (D).

**Extended data Fig. 2.**
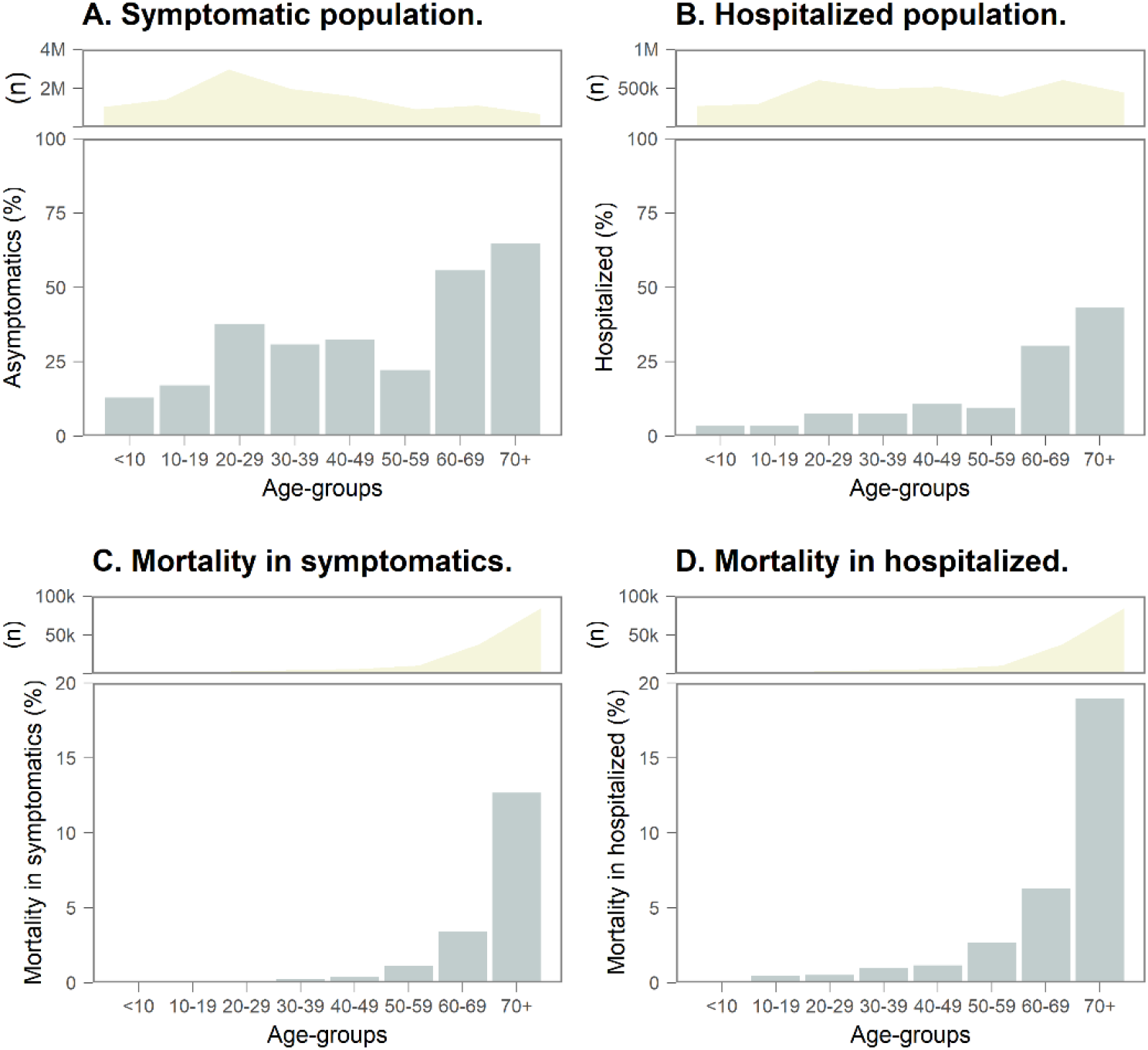
Key epidemiological outcomes from the model without social non-pharmacological interventions in Colombia.

**Extended data Fig. 3.**
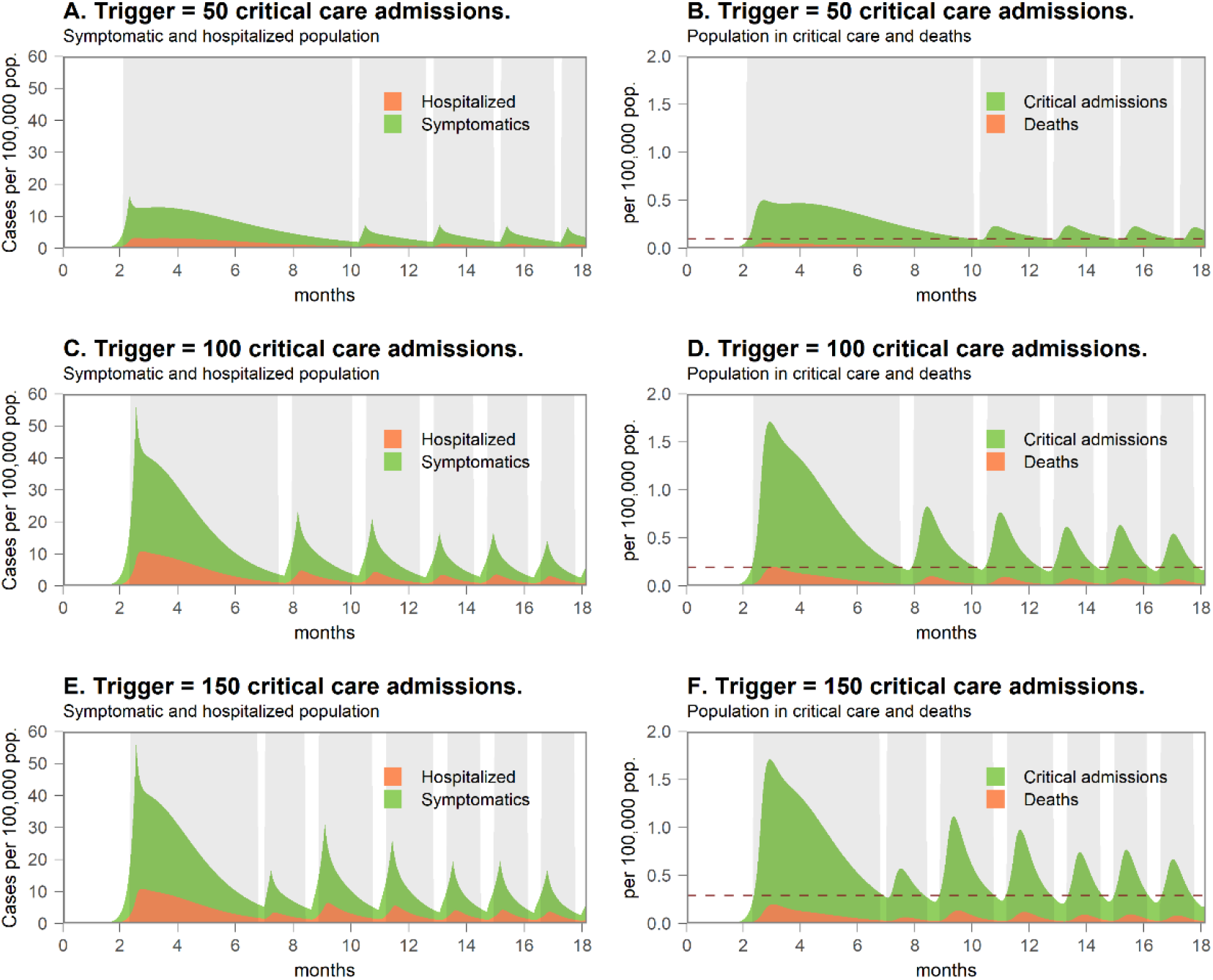
Results of including all studied social interventions (in grey area) in Colombia using different triggers (critical care admissions in a day). Note: dotted red lines in Panels B, C, and F represent the trigger levels in each of these panels.

**Extended data Fig. 4.**
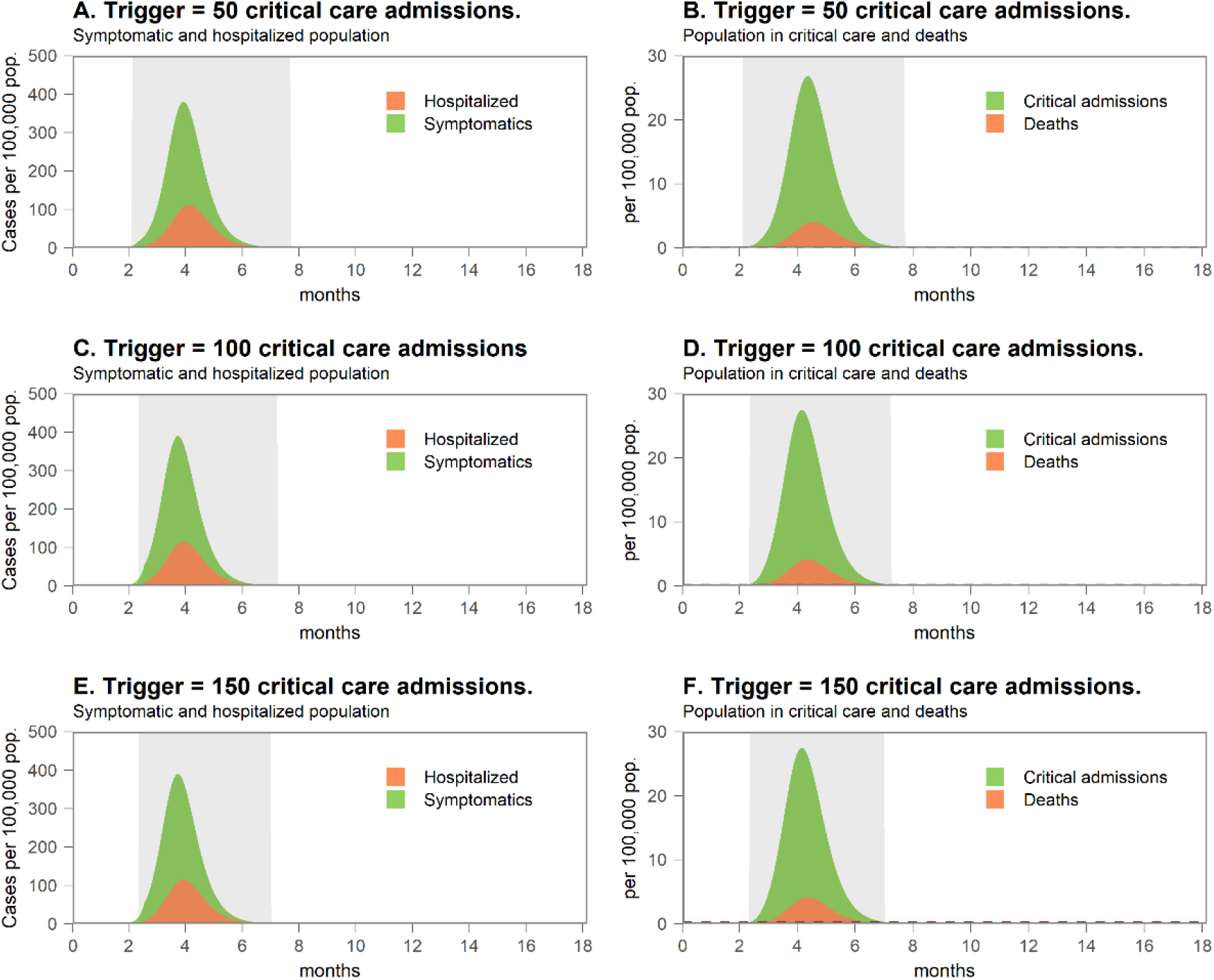
Results of including social distance (in grey area) as a non-pharmacological social intervention in Colombia using different triggers (critical care admissions in a day).

**Extended data Fig. 5.**
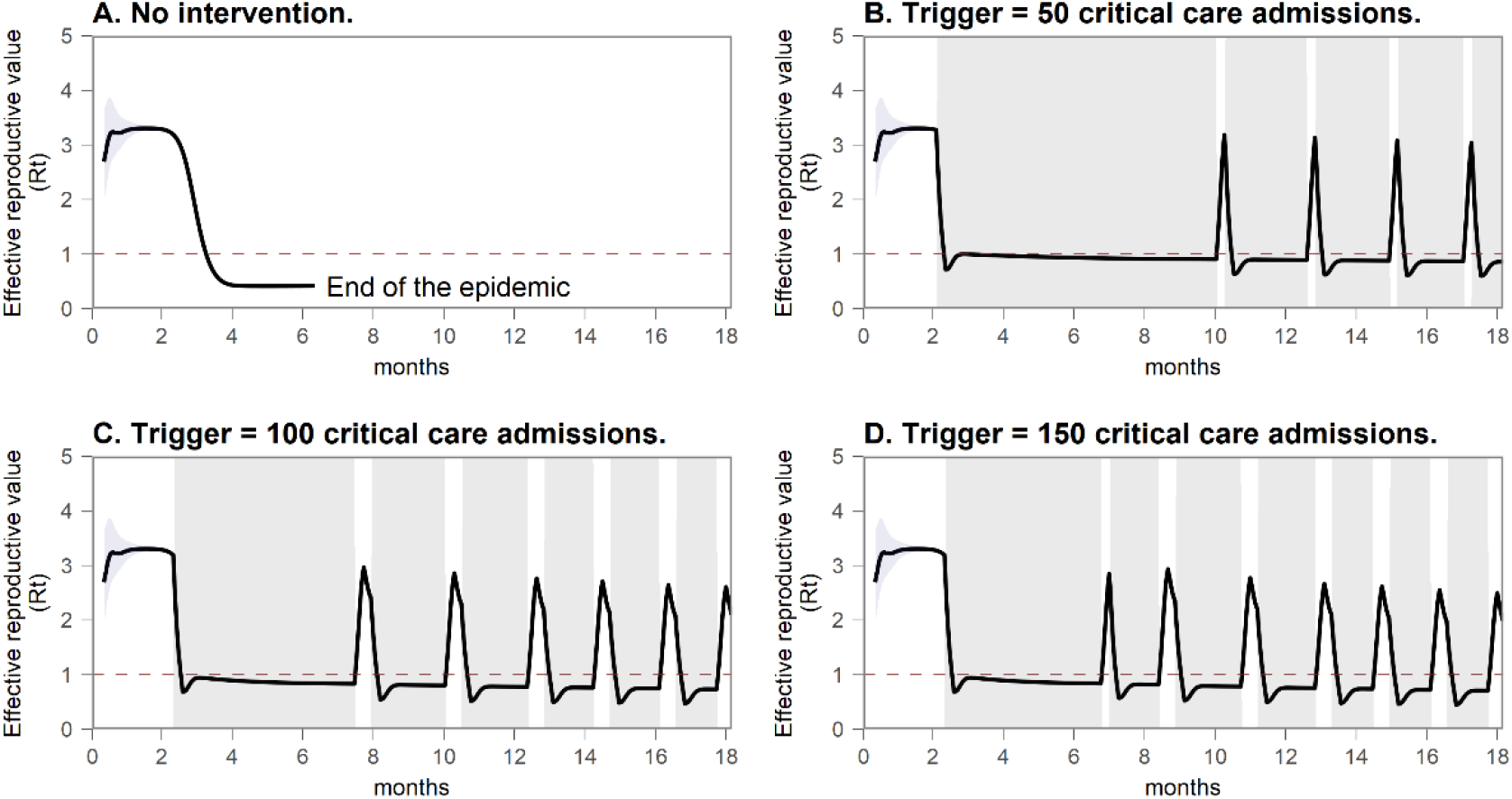
Effective reproductive value (Rt) without intervention and with all social non-pharmacological social interventions in Colombia, using different triggers.

**Extended data Fig. 6.**
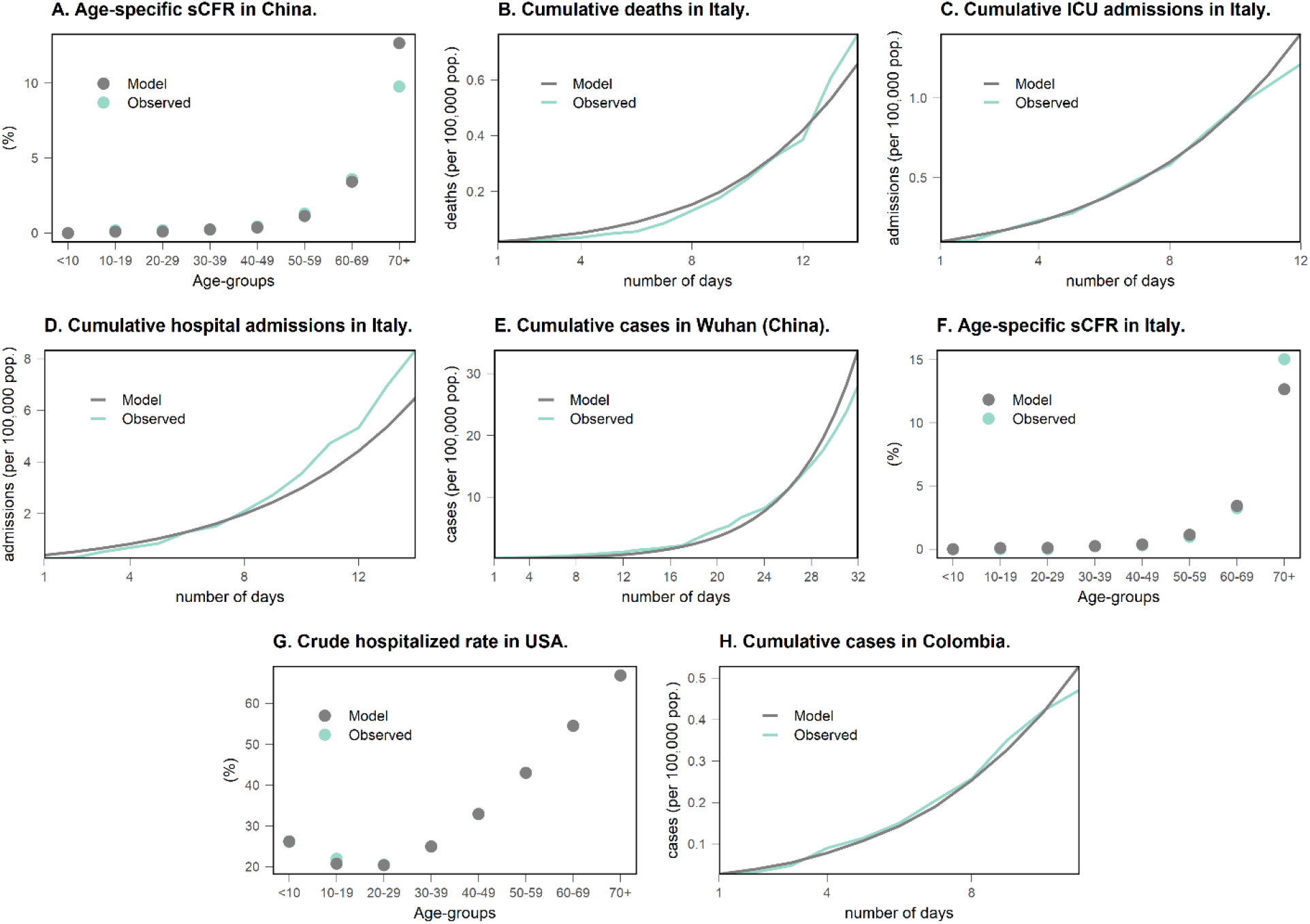
Comparison of observed data with results of best fitting values in the calibration targets. sCFR: Case-fatality rate in symptomatic population.

